# Cost-effectiveness of the second COVID-19 booster vaccination in the United States

**DOI:** 10.1101/2022.12.28.22283986

**Authors:** Rui Li, Pengyi Lu, Christopher K Fairley, José A. Pagán, Wenyi Hu, Qianqian Yang, Guihua Zhuang, Mingwang Shen, Yan Li, Lei Zhang

## Abstract

**Background:** The United States (US) authorized the second COVID-19 booster for individuals aged 50+ years on March 29, 2022. To date, the cost-effectiveness of the second booster strategy remains unassessed.

**Methods:** We developed a decision-analytic SEIR-Markov model by five age groups (0-4yrs, 5-11yrs 12-17yrs, 18-49yrs, and 50+yrs) and calibrated the model by actual mortality in each age group in the US. We conducted fives scenarios to evaluate the cost-effectiveness of the second booster strategy and incremental benefits if the strategy would expand to 18-49yrs and 12-17yrs, from a healthcare system perspective.

**Findings:** Implementing the second booster strategy for those aged 50+yrs would cost $807 million but reduce direct medical costs by $1,128 million, corresponding to a benefit-cost ratio of 1.40. Moreover, the strategy would also result in a gain of 1,048 quality-adjusted life-years (QALYs) during the 180 days, indicating it was cost-saving. Further, vaccinating individuals aged 18-49yrs with the second booster would result in an additional gain of $1,566 million and 2,276 QALYs. Similarly, expanding vaccination to individuals aged 12-17yrs would result in an additional gain of $15 million and 89 QALYs. However, if social interaction between all age groups was severed, vaccination expansion to 18-49yrs and 12-17yrs would no longer be cost-effective.

**Interpretation:** The second booster strategy was likely to be cost-effective in reducing the disease burden of the COVID-19 pandemic. Expanding the second booster strategy to 18-49yrs and 12-17yrs would remain cost-effective due to their social contacts with the older age group.

**Funding:** World Health Organization

## INTRODUCTION

Although many countries have eased COVID-19-related restrictions, the pandemic is far from over. During September 2022, about half a million new COVID-19 cases occurred every day globally.^1^ By the end of September 2022, COVID-19 has infected many more than the 0.6 billion people who were officially diagnosed with it and claimed more than 6.5 million lives.^1^ In the United States (US), over 1.1 million people have died of COVID-19 and the death toll is still rising. Effective and timely vaccination .against COVID-19 remains the best strategy for curbing the pandemic ^2-4^

A growing number of reports have raised concern that breakthrough infections are becoming increasingly prevalent.^5,6^ This is caused by both the emergence of new Omicron variants and the waning protection of COVID-19 vaccines over time.^7^ Booster shots increase the effectiveness of protection in fully vaccinated people and prevent breakthrough infections, severe conditions, or deaths due to COVID-19.^8^ As of October 1, 2022, about 100 million people or approximately one third of the US population, have received their first COVID-19 booster shots,^9^ and the number is increasing daily. This promising news, however, does not change the fact that the virus continues to mutate, and the protection provided by the booster shots continues to wane over time.^10^

A recent study demonstrated that the second BNT162b2 booster vaccine was highly effective in reducing COVID-19-related hospitalizations and deaths among older adults in Israel.^11^ Based on the available evidence on March 29, 2022, the US Food and Drug Administration (FDA) authorized “a second booster dose of the Pfizer-BioNTech COVID-19 Vaccine or Moderna COVID-19 Vaccine to be administered to individuals 50 years of age and older at least 4 months after receipt of a first booster dose of any authorized or approved COVID-19 vaccine”.^12^ Since then, millions of US adults have received their second COVID-19 vaccine booster shot. There are, however, controversies about continuing to offer a second booster shot to those who have received their first booster shot. First, offering the second booster shot would take significant resources and further increase the already enormous healthcare costs caused by the pandemic.^13^ Further, the Omicron variant as well as other newly emerged variants are less likely to cause severe conditions compared with the original SARS-CoV-2 and the earlier Alpha and Delta variants. Thus, the magnitude of the benefit of continuous vaccination against COVID-19 in the US population remains uncertain.^14^

To inform policymaking, this study assesses the cost-effectiveness of a second COVID-19 booster vaccination, which is predominately mRNA vaccines and administered 4 months after the first booster, among children, adults and older adults aged 50+ years in the US. Cost-effectiveness analysis provides important information on the trade-off between increased health benefits and increased costs associated with widely administering the second COVID-19 booster shots. Our previous study estimated the cost-effectiveness of a first COVID-19 booster vaccination among older adults in the US and concluded that the first booster shots were cost-effective.^15^ Since then, new SARS-CoV-2 variants have emerged and the proportion of the population who have been vaccinated or recovered from COVID-19 infections has changed substantially. Information on the cost-effectiveness of a second COVID-19 booster vaccination would be important for public health officials and policymakers to prioritize limited healthcare resources for continuously combating the pandemic and informing future vaccination strategies.

## METHODS

### Study design

We developed a decision-analytic SEIR-Markov model by five age groups (0-4yrs with 18,827,338 individuals, 5-11yrs with 28,584,443 individuals, 12-17yrs with 26,154,652 individuals, 18-49yrs with 138,769,369 individuals, and 50+yrs with 119,557,943 individuals) to estimate the cost-effectiveness of the second COVID-19 booster vaccination (predominately mRNA vaccines, 4 months after a first booster dose) over an evaluation period of 180 days in the United States (US). The evaluation was conducted from a healthcare system perspective. The model was constructed using TreeAge Pro 2021 R1.1, and the analysis was conducted according to the Consolidated Health Economic Evaluation Reporting Standards 2022 (CHEERS 2022) statement.^16^

### Model structure

The decision-analytic SEIR-Markov model can better capture and simulate the transmission characteristic of infectious disease as well as disease progression than a single static model.^17,18^ Existing evidence indicated that the vaccine efficacy (VE) of a first booster dose against the Omicron variant would gradually wane after four months.^12,19-21^ Thus, we defined the VE from 2 weeks to 4 months after a booster dose as a ‘short-term booster VE’, whereas the VE 4 months beyond the booster dose as a ‘long-term booster VE’.

Our model, in each age group, consisted of 13 health states including 6 uninfected states depicting varied vaccination status and 7 infected states depicting varied disease progression of COVID-19 **(Figure 1)**. People with various vaccination statuses have different risks of being infected by the Omicron strain, which was related to VEs and measured by the force of infection (λ_i,t_, i represents different age groups). The λ_i,t_ in each age group was dependent on the basic transmission coefficient and contact metrics between age groups, more details were shown in Appendix 1.1. The infected individuals first had to experience an incubation period and 31% of them would spontaneously recover without any symptoms ^22^ The remaining might first exhibit ‘mild/moderate symptoms. They might then ‘recover’ or deteriorate to a ‘severe’ state. A patient in the ‘severe’ state might ‘recover’ or progress to the ‘critical’ state. Similarly, a patient in the ‘critical’ state might ‘recover’ or ‘die’. Transition probabilities between states were estimated using the formula *p* = 1 – *e*^−*r*^, where r denoted daily transition rate.^23^ The basic model cycle length was 1 day, with a half-cycle correction applied.

**Figure 1.**
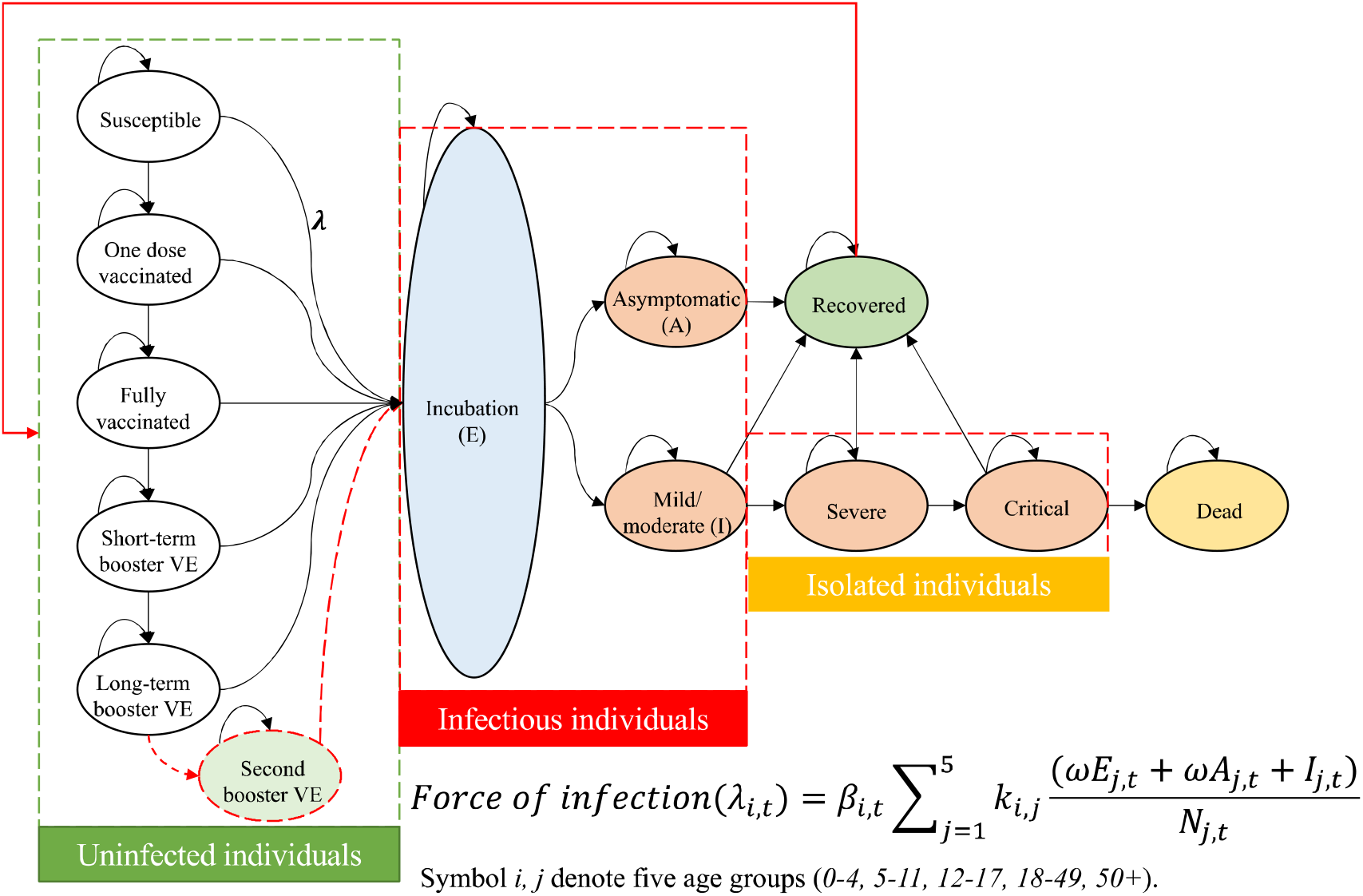
Schematic overview of the SEIR-Markov model. The *λ* _*i,t*_ the parameter of the force of infection to measure the infection risk. The symbols *i, j* denotes five age groups (0-4yrs, 5-11yrs, 12-17yrs, 18-49yrs, and 50+yrs). The *β* _*i,t*_ denotes the transmission coefficient of five age groups and the *k*_*i,j*_ represents the contact metric between age groups.

### Booster vaccine efficacy estimation

To estimate the real-world booster VE for Omicron infection and severe progression, we relied on the existing scientific literature reporting real population incidence and vaccination status data from an ongoing systematic review conducted by The International Vaccine Access Center.^24^ We obtained 99 relevant papers regarding the mRNA-based booster VE against Omicron using the version of 10^th^ Nov 2022 in this database. We screened all the studies and excluded 81 of them, of which 27 included no original data, 22 used non-unvaccinated as a reference, 10 focused on special populations, 19 studies for BA.1 Omicron, and 3 cohort studies which are not enough to produce a meta-analysis. After the exclusion of ineligible literature, we finally included 18 studies and extracted the original case data from individual studies. Then we used random-effects meta-analysis to generate the overall odds ratio (OR) and calculate the corresponding booster VE (Appendix 1.2).

### Model calibration

We refined the model inputs of transmission coefficient and vaccination rates by age groups automatically with TreeAge Pro’s calibration tool to adjust inputs until the model results match observed COVID-19-related deaths and vaccination data in the US. First, we collected the COVID-19 new deaths and vaccination data by age groups reported by the US Centres for Disease Control and Prevention (CDC).^9,25^ Then, we set the target values of the calibration as the proportion of five vaccination statuses and the accumulative deaths (day 30, 60, 90, 120, 150, 180, the simulation was started on 29^th^ March when the US approved the 2^nd^ booster) by age groups. Finally, with the input of other model parameters (**Table S1**), we used the expanded sum of square differences to measure the model goodness of fit and produced the optimal calibration results.^26^ The calibration results were shown in Appendix 1.3.

### Other model parameters

Based on the varied booster VE both for Omicron infection and for severe progression, we developed a mathematical model to estimate the distributions of clinical outcomes after being infected by Omicron in vaccinated individuals, compared to that in unvaccinated ones (Appendix 1.4). We collected the costs of booster vaccination, PCR tests and rapid antigen self-test for COVID-19 infection. In addition, we collected the cost per outpatient visit, general hospitalization and ICU admission. Then, we calculated the total direct medical cost of a COVID-19 case with varied severity by multiplying the unit cost of the medical services by the duration of each disease stage (Appendix 1.5). Health utility scores for COVID-19 patients were derived from the disutility weights of severe lower respiratory tract infection and the estimates of pricing models for COVID-19 treatments published by the Institute for Clinical and Economic Review (Appendix 1.6).

### Definition of scenarios

We defined five scenarios: Scenario 1: this counterfactual scenario assumed there was no second booster vaccination implemented in the US after first booster dose; Scenario 2: status quo, current scenario represented the actual situation of second booster vaccination for aged 50+yrs in the US, achieving coverage of 20.0% (23,927,842/119,557,943) by 24^th^ September 2022; Scenario 3: this scenario assumed all aged 50+yrs old would receive a second booster if they are eligible (4 month after first booster), this would vaccinate 23.5% more for a second booster in aged 50+yrs by 24^th^ September 2022; Scenario 4: this scenario assumed the second booster would expand for age 18-49yrs and all aged 18+yrs old would receive a second booster if they are eligible, this would achieve the coverage of 43.5% in aged 50+yrs and 26.8% in aged 18-49yrs by 24^th^ September 2022; Scenario 5: this scenario assumed the second booster would expand for age 12-49yrs and all aged 12+yrs old would receive a second booster if they are eligible, this would achieve the coverage of 43.5% in aged 50+yrs, 26.8% in aged 18-49yrs and 14.5% in aged 12-17yrs by 24^th^ September 2022. Given the vaccination coverage of the first booster in 0-4yrs and 5-11yrs was very low (0% and 4.8%, respectively), we did not include them in our scenarios for the second boosters. The presence of these two lower age groups would still facilitate the transmission of SARS-CoV-2 via social interactions.

### Model outputs

We assumed a discount rate of 3% (0-6%) annually for both cost and quality-adjusted life-years (QALYs).^27^ We calculated the costs and QALYs for the second booster vaccination strategies in each scenario and compared incremental benefits between every two consecutive scenarios of all five scenarios (scenario 2 vs. scenario 1, scenario 3 vs. scenario 2, etc.). The incremental cost-effectiveness ratio (ICER) was defined as the incremental cost per QALY gained. We used a willingness-to-pay (WTP) threshold of ICER<US$50,000.^28,29^ We also calculated the benefit-cost ratio, net monetary benefit (NMB) and cost/death saved.

### Sensitivity analysis

We conducted a univariate sensitivity analysis to examine the impact of model parameters within their respective ranges on the ICER to identify the most sensitive parameters and visualised the results using tornado diagrams. In addition, we conducted a probabilistic sensitivity analysis (PSA) based on 100,000 simulations to determine the probability of the booster strategy being cost-effective across a range of cost-effectiveness thresholds. The distributions of all model parameters were provided in Appendix 1.7.

To examine the impact of the contact matrix on the findings, we assumed the social interaction between all age groups was severed. We re-calibrated the model to the number of death cases and obtain a new contact matrix to perform sensitivity analysis (details were provided in Appendix 1.8).

## RESULTS

### The second booster strategy for those aged 50+yrs is cost-effective in the US

Compared with no second booster, vaccinating the US population aged 50+yrs with a second booster would reduce costs and increase QALYs over a period of 180 days (Table 1). Overall, the second booster strategy would incur a cost of $807 million but reduce the direct medical cost by $1,128 million, corresponding to a benefit-cost ratio of 1.40. Further, the strategy would result in a gain of 1,048 QALYs and a net monetary benefit (NMB) of $373 million over 180 days, suggesting the second booster strategy would be a cost-saving one.

**Table 1.** The results of the cost-effectiveness analysis of the second COVID-19 booster vaccination in the United States over an evaluation period of 180 days.

|  | No 2<sup>nd</sup><br>booster | Status<br>quo | Second<br>booster<br>50+ all | Second<br>booster<br>18+ all | Second<br>booster<br>12+ all | Incremental<br>benefits (SQ<br>Vs. No) | Incremental<br>benefits<br>(50+ all<br>Vs. SQ) | Incremental<br>benefits<br>(18+ all Vs.<br>50+ all) | Incremental<br>benefits<br>(12+ all Vs.<br>18+ all) |
| --- | --- | --- | --- | --- | --- | --- | --- | --- | --- |
| **QALY** | 162,321,846 | 162,322,894 | 162,326,896 | 162,329,173 | 162,329,261 | 1,048 | 4,002 | 2,276 | 89 |
| **Costs (\$, million)** | 17,186 | 16,865 | 14,157 | 12,591 | 12,576 | -321 | -2,708 | -1,566 | -15 |
| **Vaccination cost (2<sup>nd</sup> booster)** | 0 | 807 | 1,738 | 2,938 | 3,048 | 807 | 932 | 1,200 | 109 |
| 50+ yrs | 0 | 807 | 1,738 | 1,740 | 1,740 | 807 | 932 | 2 | 0 |
| 18-49 yrs | 0 | 0 | 0 | 1,198 | 1,198 | 0 | 0 | 1,198 | 0 |
| 12-17 yrs | 0 | 0 | 0 | 0 | 109 | 0 | 0 | 0 | 109 |
| **Vaccination cost (1-3 doses)** | 267 | 267 | 269 | 271 | 271 | 0 | 2 | 4 | 2 |
| **Direct medical cost** | 16,919 | 15,791 | 12,150 | 9,382 | 9,257 | -1,128 | -3,641 | -2,768 | -125 |
| 50+ yrs | 13,197 | 12,175 | 8,998 | 7,093 | 7,013 | -1,022 | -3,177 | -1,905 | -80 |
| 18-49 yrs | 2,590 | 2,516 | 2,192 | 1,535 | 1,514 | -74 | -324 | -657 | -21 |
| 12-17 yrs | 362 | 353 | 310 | 243 | 226 | -9 | -43 | -67 | -17 |
| 5-11 yrs | 362 | 352 | 308 | 244 | 240 | -10 | -44 | -64 | -4 |
| 0-4 yrs | 408 | 395 | 342 | 267 | 264 | -12 | -53 | -75 | -3 |
| **COVID-related deaths** | 65,384 | 60,135 | 44,292 | 34,236 | 33,811 | -5,249 | -15,843 | -10,056 | -425 |
| 50+ yrs | 62,761 | 57,591 | 42,109 | 32,699 | 32,299 | -5,170 | -15,483 | -9,409 | -400 |
| 18-49 yrs | 2,400 | 2,327 | 2,021 | 1,390 | 1,370 | -72 | -307 | -631 | -20 |
| 12-17 yrs | 84 | 81 | 71 | 56 | 52 | -2 | -10 | -16 | -4 |
| 5-11 yrs | 47 | 46 | 32 | 32 | 31 | -1 | -14 | 0 | -1 |
| 0-4 yrs | 92 | 89 | 60 | 60 | 59 | -3 | -29 | 0 | -1 |
| **ICER** | -- | -- | -- | -- | -- | Cost-saving | Cost-saving | Cost-saving | Cost-saving |
| **Benefit-cost ratio** | -- | -- | -- | -- | -- | 1.40 | 3.91 | 2.31 | 1.14 |
| **Cost/death prevented, \$** | -- | -- | -- | -- | -- | 153,673 | 58,818 | 119,323 | 257,184 |
| **NBM, (\$, million)** | -- | -- | -- | -- | -- | 373 | 2,908 | 1,680 | 20 |

\* Incremental benefits = difference in QALY, cost and covid-related deaths while two scenarios comparing.
Benefit-cost ratio: the ratio between the reduction in the direct medical cost for COVID-19 disease and the investment in 2<sup>nd</sup> booster vaccination
Net monetary benefit (NMB) is calculated as (incremental benefit × willingness-to-pay threshold) – incremental cost.

Second booster vaccination would prevent 848,584 new COVID-19 infections (62% in the 50+yrs group, 29% in the 18-49yrs group), 41,806 hospitalizations (95% in 50+yrs group, 3% in the 18-49yrs group), 7,267 ICU-admissions (94% in 50+yrs group, 4% in 18-49yrs group), 5,249 COVID deaths (99% in 50+yrs group, 1% in 18-49yrs group), indicating a requirement of $153,673 to prevent one COVID death (Figure 2).

**Figure 2.**
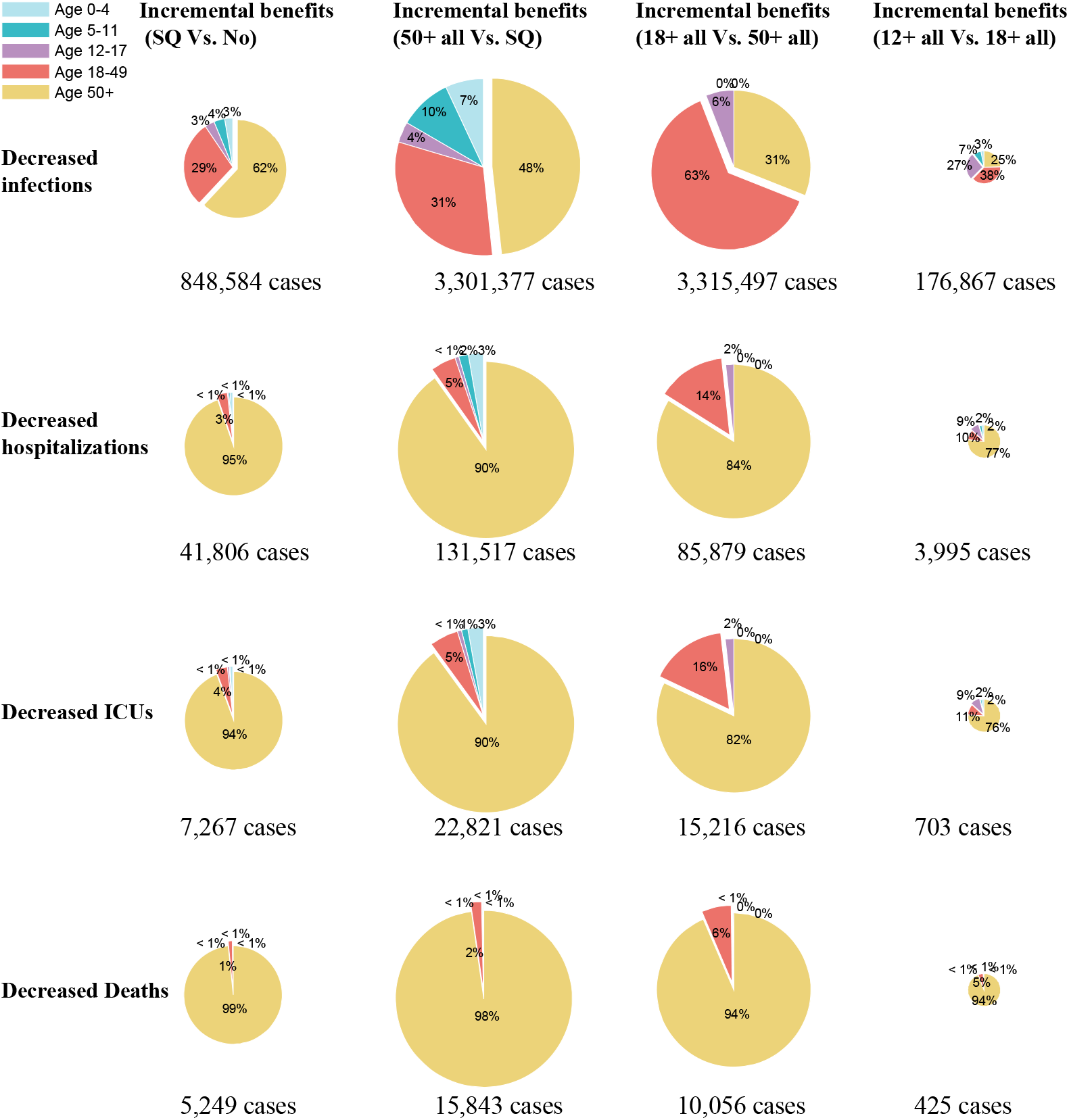
The incremental benefits of decreased COVID-19 infections and severe clinical outcomes of the second COVID-19 booster strategy in varied scenarios in the United States. Abbreviations: Scenario 1: No 2^nd^ booster strategy, No; Scenario 2: Status quo, SQ; Scenario 3: 2^nd^ booster strategy for all eligible 50+ individuals, 50+all; Scenario 4: 2^nd^ booster strategy for all eligible 18+ individuals, 18+all; Scenario 5: 2^nd^ booster strategy for all eligible 12+ individuals, 12+all.

Univariate sensitivity analysis showed that varying most of the model parameters individually would not alter the conclusion of the cost-effectiveness of the second booster. The top three most sensitive parameters on ICER were transmissibility of the Omicron variant, direct medical cost and vaccination cost, and VE for Omicron infection (Figure 3A). Probabilistic sensitivity analysis (PSA) based on 100,000 simulations demonstrated the probability of being cost-effective with the second booster strategy was 67.58% (Figure 4A).

**Figure 3.**
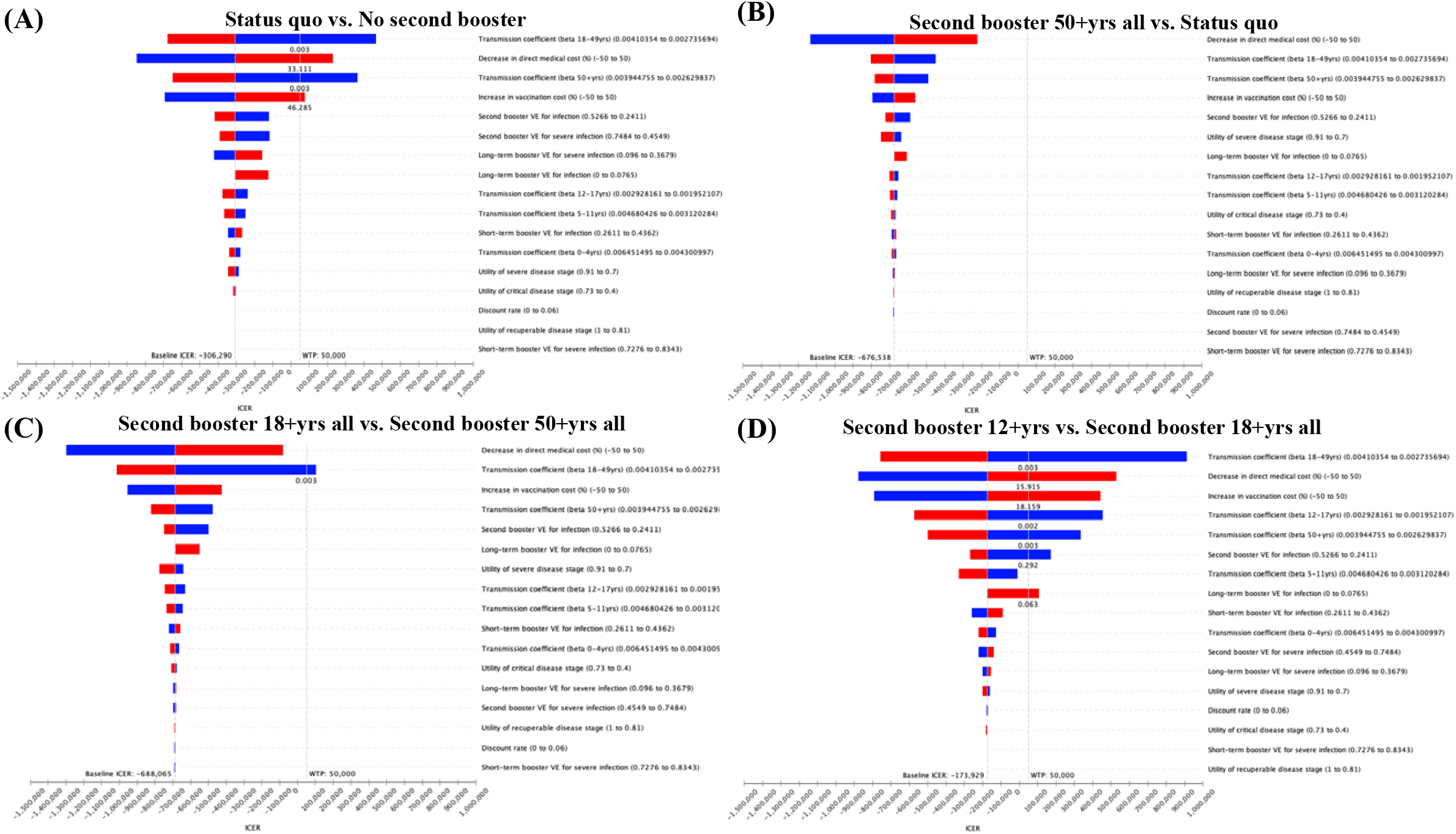
The tornado plot of one-way sensitivity analyses of the second COVID-19 booster vaccination strategy implemented in different age groups in the United States. A horizontal bar was generated for each parameter analysis. The width of the bar indicates the potential effect of the associated parameter on the ICER when the parameter is changed within its range. The red part of each bar indicates high values of input parameter ranges, while the blue part indicates low values. The dotted vertical line represents the threshold of willingness-to-pay (WTP) of the baseline.

**Figure 4.**
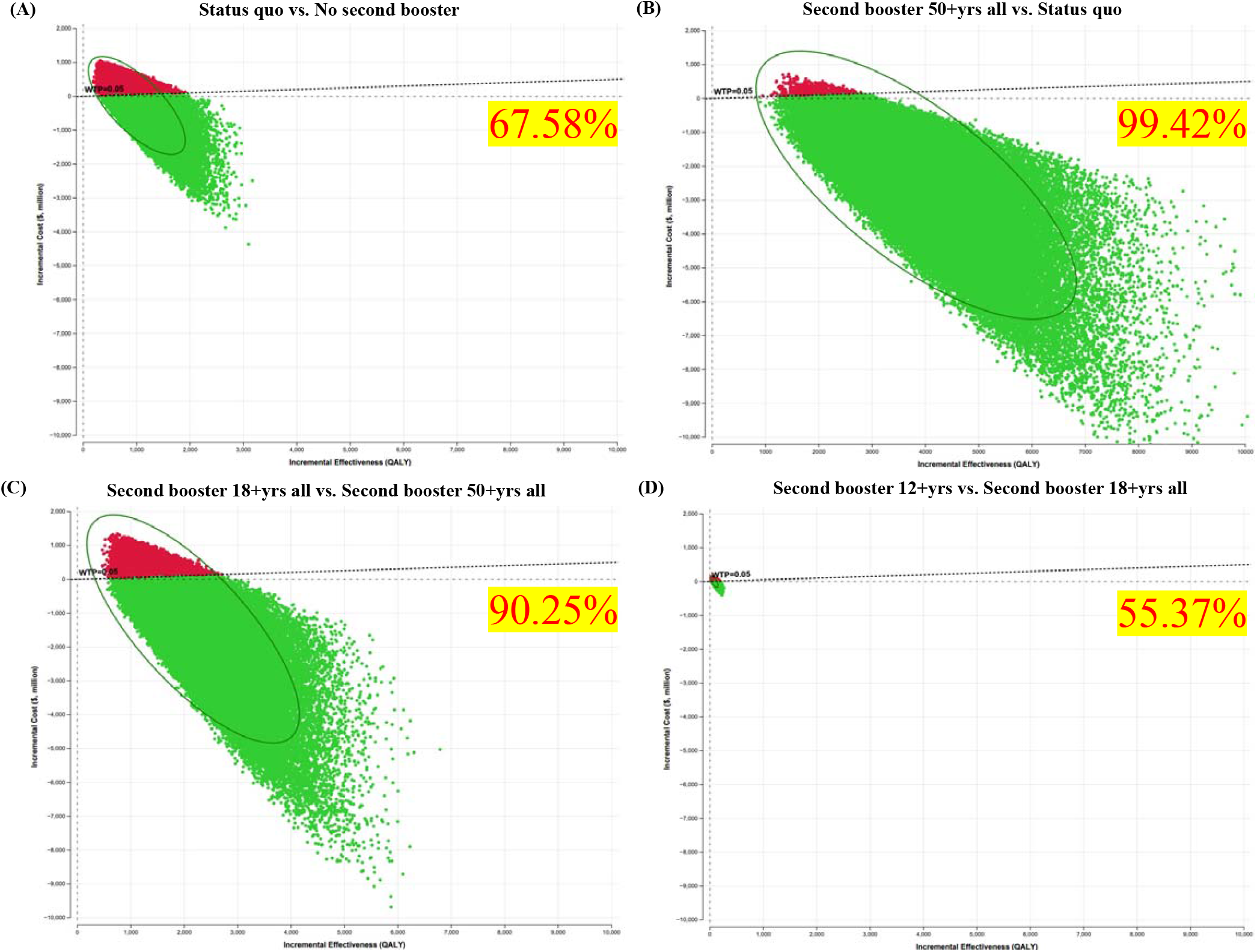
Probabilistic sensitivity analysis (PSA) based on 100,000 simulations (the figures mean the chance of being cost-effective) of the second COVID-19 booster vaccination strategy implemented in different age groups in the United States.

### Incremental benefits of the second booster expansion to all eligible 50+yrs individuals

Compared with the status quo, vaccinating all eligible 50+yrs with a second booster (second booster 50+yrs all scenario) would further result in a reduced cost of $2,708 million, consisting of the increased vaccination cost of $932 million and reduced direct medical care cost of $3,641 million. This corresponded to a benefit-cost ratio of 3.91. Further, the strategy would result in a gain of 4,002 QALYs and an NMB of $2,908 million during the 180 days (Table 1). Moreover, the strategy would reduce 3.3 million infections (48% in the 50+yrs group, 31% in the 18-49yrs group), 131,517 hospitalizations (90% in the 50+yrs group, 5% in 18-49yrs group), 22,821 ICU-admissions (90% in 50+yrs group, 5% in 18-49yrs group), 15,843 COVID deaths (98% in 50+yrs group, 2% in 18-49yrs group), indicating a cost of $58,818 to prevent one COVID death (Figure 2). The univariate sensitivity analysis showed that varying any individual model parameter at one time would not change the conclusion of the cost-effectiveness of the strategy (Figure 3B). The probabilistic sensitivity analysis demonstrated a very high probability (99.42%) of this strategy being cost-effective (Figure 4B).

### Incremental benefits of expanding the second booster for all eligible 18-49yrs age group

Compared with the second booster 50+yrs all scenario, vaccinating all eligible 18+yrs individuals (second booster 18+ all scenario) would further result in a reduced cost of $1,566 million (1,200 million increased vaccination cost and 2,768 million reduced direct medical cost) and a gain of 2,276 QALYs, corresponding to a benefit-cost ratio of 2.31 and an NMB of $1,680 million (Table 1). Likewise, the strategy would reduce 3.3 million infections and most of the prevented infections (63%) are in the 18-49yrs age group. Surprisingly, most of the prevented hospitalizations (84%), ICU admissions (82%), and deaths (94%) of this strategy are aged 50+yrs. This indicates most of the incremental benefits of this strategy were attributed to the decreased burden in those aged 50+ years (Figure 2). The sensitivity analyses also showed stable results for this strategy to be cost-effective (Figure 3C, 4C).

### Incremental benefits of expanding the second booster for all eligible 12-17yrs age group

Compared with the second booster 18+yrs all scenario, vaccinating all eligible 12+yrs individuals would further result in a reduced cost of $15 million (109 million increased vaccination cost and 125 million reduced direct medical cost) and a gain of 89 QALYs, corresponding to a benefit-cost ratio of 1.14 and an NMB of $20 million (Table 1). Similarly, most of the prevented hospitalizations (77%), ICU admissions (76%), and deaths (94%) of this strategy are in the 50+ years age group (Figure 2). The univariate sensitivity analysis showed that varying most individual model parameters at one time could not change the conclusion of the cost-effectiveness of the strategy (Figure 3D). The probabilistic sensitivity analysis demonstrated a probability of 55.37% for this strategy to be cost-effective (Figure 4D).

### Impact of social contacts between age groups on second booster cost-effectiveness

To examine the impact of the contact matrix on the study findings, we arbitrarily severed all social contacts between different age groups and re-calibrated the model (Appendix 1.8). In this scenario, providing a second booster shot to those aged 50+yrs (status quo and second booster 50+ all scenario) would result in more gain of QALYs than that in an unsevered situation in the context of the same epidemic scale. Moreover, the second booster strategy would save more direct medical costs with a high benefit-cost ratio of 3.00 and 7.30, respectively (Table S2). In contrast, applying a second booster for those aged 18-49yrs and 12-17yrs (second booster 18+ all scenario and second booster 12+ all scenario) is no longer cost-effective with an ICER value of $754,345 and $859,594/QALY gained, respectively.

## DISCUSSION

Our study evaluated the population impacts and cost-effectiveness of the second booster vaccination strategy in the US population. We identified several key findings. First, compared to no second booster strategy, the second booster strategy would reduce 1.3 million infections, 41,806 hospitalizations, 7,269 ICU-admission, and 5,249 deaths among adults aged 50 years and older. This would reduce direct medical costs by $1,128 million but only incur an additional vaccination cost of $807 million, corresponding to a benefit-cost ratio of 1.40. Moreover, the strategy would result in a gain of 1,041 QALYs during 180 days, suggesting the second booster strategy was cost-saving. Likewise, it is more cost-effective if all eligible individuals aged 50 years and older received their second boosters. Notably, if we expand the second booster strategy to those aged 18-49yrs, it will reduce direct medical costs by $2,768 million and the majority of decreased costs were attributed to the 50+yrs age group ($1,905 million). This scenario incurs an additional vaccination cost of $1200 million and a gain of 2,276 QALYs, leading to an overall cost-saving result. Similarly, further expansion to the 12-17yrs would also be cost-saving. However, if the-infection only transmits within each age group, the expansion 18-49yrs and 12-17yrs would no longer be cost-effective. This indicates the second booster strategy for 18-49yrs and 12-17yrs would remain cost-saving when their interaction with other (elder) age groups was explicitly modelled but not cost-effective if the interaction was severed.

Our study demonstrates that the second booster for those aged 50+ years is likely to be cost-effective during the Omicron pandemic in the US, and it would be more cost-effective if all eligible 50+yrs individuals received a second booster. Even though the Omicron strain is less pathogenic than delta and even other previous SARS-COV-2 strains, it still causes a large disease burden in the US, mostly from the older age group.^25^ Currently, nearly 98% of deaths are in people 50 or older in the US and, thus, it is crucial to reinforce the preventive and treatment strategies to protect the elder people. A second booster shot could not only improve protection against Omicron both for preventing infection and progression to severe disease status,^30^ but also reduce billions of direct medical costs due to the reduced number of severe patients. Promoting taking the second booster dose for those aged 50+ should be still the priority to suppress the Omicron epidemic in the elderly.

Our study demonstrates that expanding the second booster strategy for those aged 12-49yrs is likely to be cost-effective, largely because of reducing the disease burden and medical costs in those aged 50+ years. Compared to the 1.067% of fatality rate in aged 50+yrs COVID-19 infections, individuals aged 18-49, and 12-17 have milder symptoms and lower fatalities of 0.033% and 0.009%, respectively.^25,31^ Although the disease burden and health impact in the younger age groups is mild *per se*, our study found an unexpected increase in the benefit for the elderly by enhancing vaccination in the younger population. Providing a second booster for individuals aged 18-49yrs adults bring huge incremental benefits for those aged 50+yrs, and it would reduce 3.3 million Omicron infections (31% in 50+yrs), 85,879 hospitalizations (84%), 10,056 deaths (94%), and 2,758 million medical costs (69%) deaths in 180 days in the US. This finding provides a new perspective to evaluate the additional value of the second booster strategy to suppress the pandemic.

Our study demonstrates that the second booster strategy for 18-49yrs and 12-17yrs alone would not be cost-effective for curbing the Omicron epidemic if they do not interact with other age groups. Assuming all the 18-49yrs and 12-17yrs people interact only with their peers but total contacts remain consistent with the previous epidemic scale, the strategies would no longer be cost-effective. This is mainly due to mild symptoms after Omicron infection in younger age groups and most of them would spontaneously recover without any medical treatments. In this situation, the Omicron epidemic would not bring a huge disease burden and medical costs for those aged 18-49yrs or 12-17yrs, and thus it would be less cost-effective to receive a second booster shot for their own.

Our study has several limitations. First, although we used an SEIR-Markov model to simulate the COVID-19 epidemic, our model still lacks the ability to predict future epidemics and new variants, and thus our findings should be interpreted with all other resources available when informing public health initiatives for vaccination and epidemic management. Second, based on the US recommendation schedule for second booster vaccination, we assumed that the VE of the first booster wanes 4 months after vaccination. In reality, the efficacy of vaccines is likely to gradually decline without a clear cut-off. This assumption may have led to an overestimate of booster VE in the short term, and an underestimate of booster VE in the long term. Third, we assumed the immune individuals would lose the COVID-19 infection-induced antibody protection and reverse back to the un-infected health states in the model so that it simulates the fact the infected individuals can be reinfected again. However, the transition probability of losing the protection from the previous infection remained uncertain and estimated by our model calibration as 0.033% per day, which means about 11% of the immune individuals would reverse back to the uninfected states annually. Finally, we only consider the mRNA-based COVID-19 vaccines (BNT162b2, mRNA1273) in our model. Nevertheless, in reality, mRNA-based COVID-19 vaccines are most used in the US and account for 96.8% of vaccination use.^9^

In conclusion, the second booster strategy implemented in the US is likely to be beneficial and cost-effective for reducing the disease burden of the COVID-19 pandemic. Expanding the second booster strategy to 18-49yrs and 12-17yrs would remain cost-effective due to their contacts with the elderly age groups.

## Data Availability

All data produced in the present work are contained in the manuscript

## Declarations

### Ethics approval and consent to participate

Not applicable

### Competing interests

The authors declare that they have no competing interests.

### Data sharing

All data relevant to the study are included in the Article or in the online appendix.

### Authors’ contributions

RL and LZ conceived the study. RL was involved in the study concept and design, data acquisition, data analysis, interpretation of data, and drafting of the manuscript. PL and QY were involved in the acquisition of data. CF, JP, WH and GZ were involved in the critical revision of the manuscript. LZ, YL and MS were involved in overall study supervision. RL, LZ, YL and MS had access to and verified the data. All authors participated in preparing the manuscript and have seen and approved the final version for submission.

## Acknowledgments

This study is supported by the World Health Organization (Grant number: 2022/1262416-0); LZ is supported by the Bill & Melinda Gates Foundation (Grant number: INV-006104); the National Natural Science Foundation of China (Grant number: 81950410639); Outstanding Young Scholars Support Program (Grant number: 3111500001); Xi’an Jiaotong University Basic Research and Profession Grant (Grant number: xtr022019003, xzy032020032); Epidemiology modelling and risk assessment (Grant number: 20200344) and Xi’an Jiaotong University Young Scholar Support Grant (Grant number: YX6J004).

## Notes

### Competing Interest Statement

The authors have declared no competing interest.

### Funding Statement

LZ is supported by the National Natural Science Foundation of China (Grant number: 81950410639), Outstanding Young Scholars Funding (Grant number: 3111500001), Xi'an Jiaotong University Basic Research and Profession Grant (Grant number: xtr022019003 and xzy032020032), and Xi'an Jiaotong University Young Talent Support Grant (Grant number: YX6J004). We are grateful to Kiyohiko Izumi for his skilful review of the published work.

### Summary of Updates

Updated version of manuscript

