## Appendix for "Cost-effectiveness of the second COVID-19 booster vaccination in the United States"

This supplementary document describes in detail the model construction and estimation of parameters presented in the main text.

#### 1.1 Health states and the force of infections

We developed a decision-analytic SEIR-Markov model by five age groups (0-4yrs with 18,827,338 individuals, 5-11yrs with 28,584,443 individuals, 12-17yrs with 26,154,652 individuals, 18-49yrs with 138,769,369 individuals, and 50+yrs with 119,557,943 individuals) to simulate the disease transmission and progression of the Omicron variant over a period of 180 days in the United States (US). Our model, in each age group, consisted of 13 health states including 6 uninfected states depicting varied vaccination status and 7 infected states depicting varied disease progression of COVID-19 **(Figure 1).**

We defined the 13 health states as follows.

- Susceptible: individuals who have not been vaccinated.
- One dose vaccinated: individuals who have revived one dose vaccination.
- Fully vaccinated: individuals who have completed series vaccinations.
- Short-term booster VE: the vaccine efficacy from 2 weeks to 4 months after a booster dose.
- Long-term booster VE: the vaccine efficacy 4 months beyond a booster dose.
- Second booster VE: the vaccine efficacy after a second booster shot.
- Incubation (E): cases prior to symptom onset.
- Asymptomatic (A): cases who never developed symptoms ever throughout the course of their disease.
- Mild/moderate (I): cases without pneumonia and cases with mild pneumonia.
- Severe: cases who developed dyspnoea and/or hypoxemia and managed in a hospital but not requiring intensive care unit.
- Critical: cases who developed respiratory failure, and/or septic shock, and/or multiple organ dysfunction/failure and managed in an intensive care unit; some of them who recuperated from critical disease need to go through the recuperation stage-remaining in the hospital or other health care facility
- Recovered: cases who recovered from infection stages.
- Death: COVID-19 related death

People with various vaccination status by age group have different risk of being infected and different disease progression to different clinical outcomes. For the susceptible individuals, the infection risk is measured by the parameter of force of infection ($\lambda_{i,t}$), $i,j$ denotes five age groups (0-4yrs, 5-11yrs, 12-17yrs, 18-49yrs, and 50+yrs). The $\lambda_{i,t}$ is given by

$$\lambda_{i,t} = \beta_{i,t} \sum_{j=1}^{5} k_{i,j} \frac{(\omega E_{j,t} + \omega A_{j,t} + I_{j,t})}{N_{i,j}}$$

Where the transmission coefficient ($\beta_{i,t}$) denotes the probability that a susceptible individual has been infected by contacting other infectious cases by age group once. The $\lambda_{i,t}$ is determined by the contact metric between age groups ($k_{i,j}$) and number of infectious patients (E, A, I). Previous studies had reported a 75% lower infectiveness (1-$\omega$) of latent, asymptomatic individuals (E, A) compared with symptomatic individuals (I)[1]. The contact metric in the US is obtained from the polished literature as follows. [2]

| Susceptible \ Infections | 0-4 | 5-11 | 12-17 | 18-49 | ≥50 |
|---|---|---|---|---|---|
| 0-4 | 2.24 | 1.32 | 0.67 | 5.02 | 2.40 |
| 5-11 | 1.08 | 6.19 | 2.61 | 5.16 | 2.24 |
| 12-17 | 0.40 | 3.99 | 7.27 | 7.41 | 2.06 |
| 18-49 | 0.60 | 1.00 | 1.46 | 8.60 | 2.76 |
| ≥50 | 0.38 | 0.60 | 0.59 | 4.97 | 3.99 |

#### 1.2 Real-world vaccine efficacy against Omicron infection and severe progression

To identify real-world vaccine efficacy ( VE) against Omicron infection and severe progression, we collected multiple scientific literature including real population incidence among varied vaccination status groups from an ongoing systematic review conducted by The International Vaccine Access Center[3]. We searched the published report version (10th Nov 2022) and found 99 relevant papers regarding the mRNA-based booster vaccine effectiveness against Omicron (80 booster VE papers and 26 second booster VE papers, of which 7 repeated). We scrutinized the full texts of all papers for eligibility, of which 81 were excluded (27 studies included no original cases number; 22 studies adopt vaccinated individuals as reference; 10 studies were focused on special population such as pregnancies, previous infection and patients receiving haemodialysis; 19 studies included sample during BA. 1 Omicron dominant period; 3 studies were designed by cohort research and not enough to produce meta-analysis). After elimination of ineligible literature, we finally included 18 studies from an ongoing systematic review (17 booster VE papers and 5 second booster VE papers, of which 4 repeated).

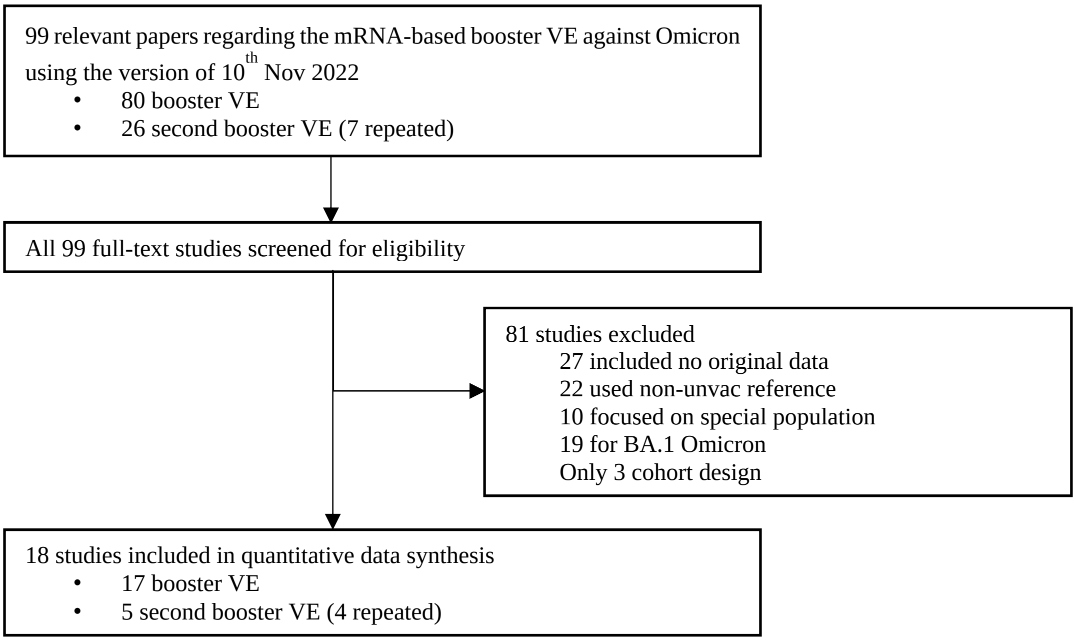


Existing evidence indicated that the booster VE would gradually wane with time and the Centre of Disease Control and Prevention (CDC) recommended second booster for those aged ≥50 years 4 months after their prior dose to increase their protection further[4-7]. Thus, we defined the VE from 2 weeks to 4 months after a booster dose as a 'short-term booster VE', whereas the VE beyond 4 months as a 'long-term booster VE'. Combined with the VE classification of preventing Omicron infection and severe progression, we extracted original cases data using standard tables designed by case-control studies (details were shown in Additional file.xlsx). Then, we used random-effects meta-analysis to generate overall odds ratio (OR) estimates. Finally, we calculated the VE using the formula of (1–OR) multiplied by 100%.[5]

| **Variables** | **Pooled OR** | **VE** (%) |
| --- | --- | --- |
| Short-term booster VE for preventing non-BA.1 infection | 0.645(0.739, 0.564) | 35.5 (26.1, 43.6) |
| Short-term booster VE for preventing severe disease | 0.212 (0.272, 0.166) | 78.8 (72.8, 83.4) |
| Long-term booster VE for preventing non-BA.1 infection | 1.268 (1.740, 0.924) | -26.8 (-74.0, 7.7) |
| Loge-term booster VE for preventing severe disease | 0.756 (0.904, 0.632) | 24.4 (9.6, 36.8) |
| 2nd booster VE for preventing non-BA.1 infection, compared with no vaccination | 0.599 (0.759, 0.473) | 40.1 (24.1, 52.7) |
| 2nd booster VE for preventing severe disease, compared with no vaccination | 0.370 (0.545, 0.252) | 63.0 (45.5, 74.8) |

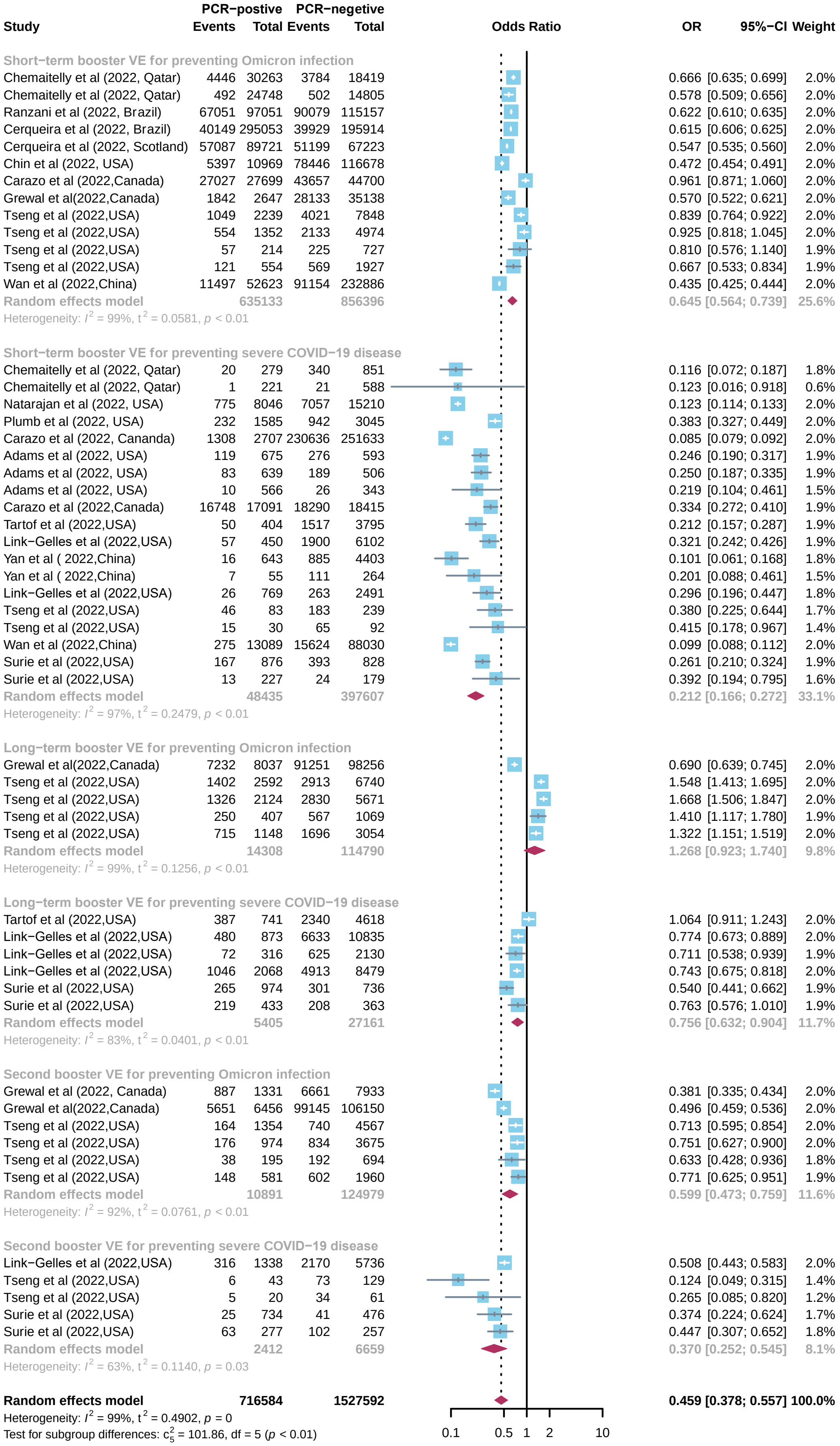


**Figure S1**. Forest plot for the pooled odds ratio for Omicron variant.

#### 1.3 Model calibration

We refined the model inputs of transmission coefficient and vaccination rates by age groups automatically with TreeAge Pro's calibration tool to adjust inputs until the model results match observed COVID-19 related mortality and vaccination data in the US. The model calibration results are shown as follow.

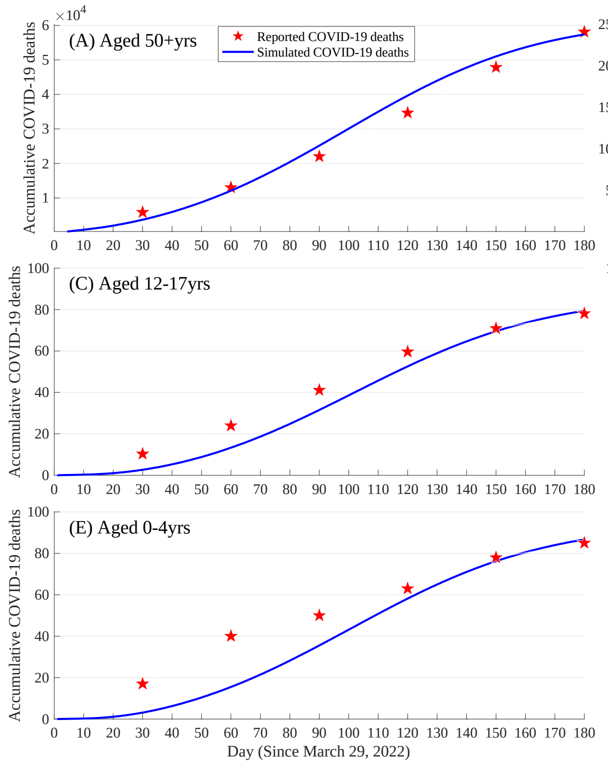


**Figure S2.** Model calibration for COVID-19 related deaths by age groups in the US.

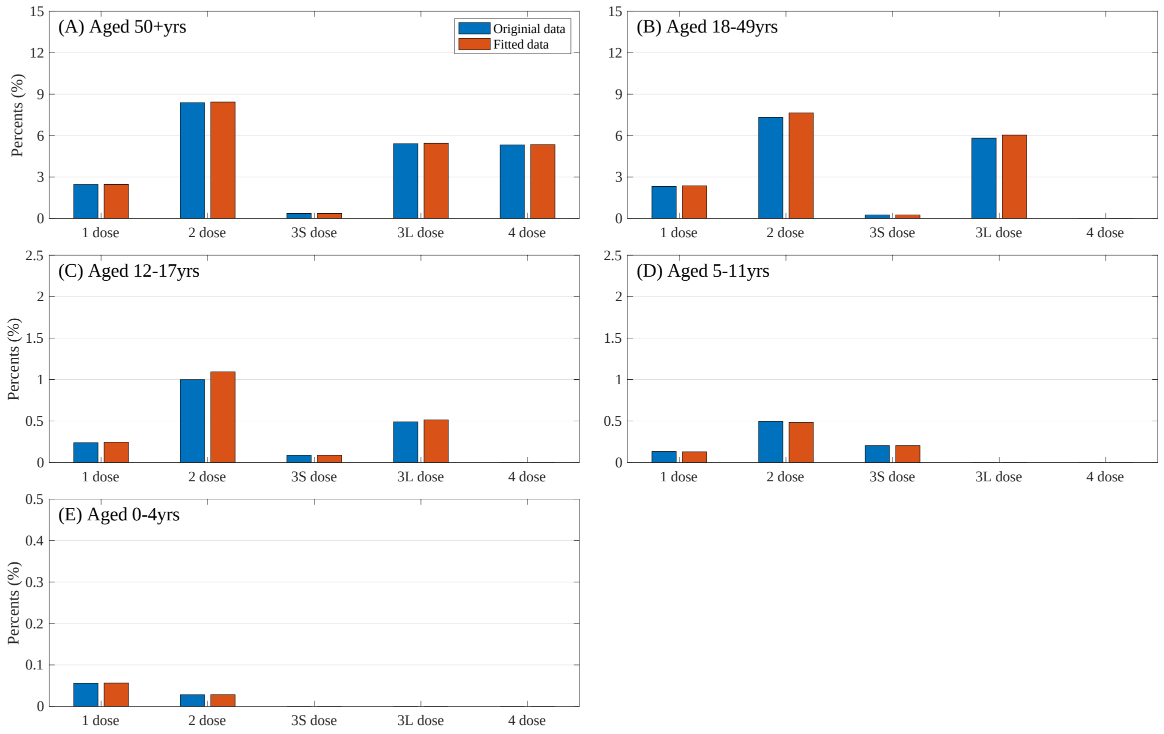


**Figure S3.** Model calibration for vaccination status of uninfected individuals by age groups in the US.

The calibrated parameters are shown as follows:

| Parameters | Description | Values |
|---|---|---|
| $\beta_{1,0}$ | The basic transmission coefficient of those aged 0-4yrs | 0.005376 |
| $\beta_{2,0}$ | The basic transmission coefficient of those aged 5-11yrs | 0.003900 |
| $\beta_{3,0}$ | The basic transmission coefficient of those aged 12-17yrs | 0.002440 |
| $\beta_{4,0}$ | The basic transmission coefficient of those aged 18-49yrs | 0.003420 |
| $\beta_{5,0}$ | The basic transmission coefficient of those aged 50+yrs | 0.003287 |
| $a$ | The amplitude of the function $\beta_{i,t} = \beta_{i,0} * a * (2 + sin\ (ct + b))$ | 2.866473 |
| $b$ | The phase shift of the function $\beta_{i,t} = \beta_{i,0} * a * (2 + sin\ (ct + b))$ | 2.422840 |
| $c$ | The period of function $\beta_{i,t} = \beta_{i,0} * a * (2 + sin\ (ct + b))$ | 0.010000 |
| $V_{lose}$ | The transition probability of protection loses | 0.000333 |
| $Vac_{11}$ | The vaccination rate of first dose of those aged 0-4yrs | 0.000350 |
| $Vac_{12}$ | The vaccination rate of second dose of those aged 0-4yrs | 0.004990 |
| $Vac_{21}$ | The vaccination rate of first dose of those aged 5-11yrs | 0.000036 |
| $Vac_{22}$ | The vaccination rate of second dose of those aged 5-11yrs | 0.001359 |
| $Vac_{23}$ | The vaccination rate of first booster dose of those aged 5-11yrs | 0.002138 |
| $Vac_{31}$ | The vaccination rate of first dose of those aged 12-17yrs | 1.09E-08 |
| $Vac_{32}$ | The vaccination rate of second dose of those aged 12-17yrs | 0.000319 |
| $Vac_{33}$ | The vaccination rate of first booster dose of those aged 12-17yrs | 0.000623 |
| $Vac_{34}$ | The transition probability of short-term booster to long-term of those aged 12-17yrs | 0.014935 |
| $Vac_{41}$ | The vaccination rate of first dose of those aged 18-49yrs | 8.38E-15 |
| $Vac_{42}$ | The vaccination rate of second dose of those aged 18-49yrs | 0.000081 |
| $Vac_{43}$ | The vaccination rate of first booster dose of those aged 18-49yrs | 0.000364 |
| $Vac_{44}$ | The transition probability of short-term booster to long-term of those aged 18-49yrs | 0.021638 |
| $Vac_{51}$ | The vaccination rate of first dose of those aged 50+yrs | 0.000114 |
| $Vac_{52}$ | The vaccination rate of second dose of those aged 50+yrs | 0.000456 |
| $Vac_{53}$ | The vaccination rate of first booster dose of those aged 50+yrs | 0.000647 |
| $Vac_{54}$ | The transition probability of short-term booster to long-term of those aged 50+yrs | 0.004403 |
| $Vac_{55}$ | The vaccination rate of second booster dose of those aged 50+yrs | 0.021681 |

#### 1.4 Distribution of clinical disease stages of various vaccination status

We collected the distribution of clinical disease stages of unvaccinated individuals (**Group A**) infected by Omicron variant by age groups from the Centers for Disease Control and Prevention (CDC) in the US and published literature.

| Mar 29, 2022 to Sep 24, 2022 | | Cumulative Case[8] | Cumulative Death[9] | Fatality (Die% in infections) | ICU% in infections | Hos% in infections | ICU% in Hos | Die% in Hos |
|---|---|---|---|---|---|---|---|---|
| **1** | All Ages | 15289158 | 60670 | **0.397%** | | | | |
| **2** | 0-4yrs | 663426 | 85 | **0.013%** | **0.281%** | **1.55%[10]** | 18.11%[10,11] | 0.83% |
| **3** | 5-11yrs | 966299 | 44 | **0.005%** | **0.085%** | **0.77%[10]** | 11.08%[12] | 0.59% |
| **4** | 12-17yrs | 908620 | 78 | **0.009%** | **0.147%** | **0.77%[10]** | 19.05%[13] | 1.12% |
| **5** | 18-49yrs | 7304308 | 2375 | **0.033%** | **0.127%** | **0.64%** | 19.93%[10] | 5.10%[14] |
| **6** | 50yrsmore | 5446505 | 58113 | **1.067%** | **1.415%** | **8.18%** | 17.31%[10] | 13.05%[14] |

Based on the short-term booster VE (**Group B**), long-term booster VE (**Group C**) and 2nd booster VE (**Group D**), we developed a mathematical model to estimate the similar distributions in varied vaccination group (**Figure S4**). In the model, we calculated the numerator and denominator of the proportion of severe infections in vaccinated patients ($P_{vs}$) by three factors, which are the proportion of severe infections in unvaccinated infections ($P_s$), the VE for preventing Omicron infection ($V_i$) and severe progression ($V_s$). Then, we could calculate the respective proportion of clinical outcomes by vaccination status.

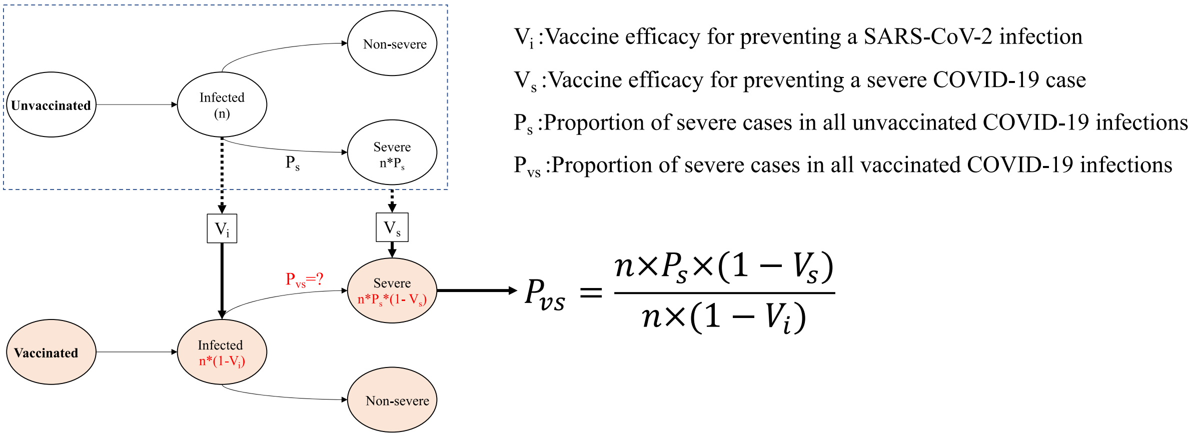


**Figure S3**. The model flowchart to estimate the proportion of severe cases in the COVID-19 infections.

The details of the distribution are list as follow.

| **Clinical outcomes** | **Group A_50+yrs** | **Group B_50+yrs** | **Group C_50+yrs** | **Group D_50+yrs** | **Group A_18-49yrs** | **Group B_18-49yrs** | **Group C_18-49yrs** | **Group D_18-49yrs** |
|---|---|---|---|---|---|---|---|---|
| Asymptomatic infection | 31.00% | 32.85% | 31.67% | 32.05% | 31.00% | 33.50% | 33.41% | 33.44% |
| Mild/Moderate illness | 60.82% | 64.46% | 62.15% | 62.89% | 68.36% | 66.29% | 66.11% | 66.17% |
| Severe illness | 6.760% | 2.224% | 5.110% | 4.177% | 0.511% | 0.168% | 0.386% | 0.316% |
| Critical (recover) illness | 0.348% | 0.115% | 0.263% | 0.215% | 0.095% | 0.031% | 0.072% | 0.058% |
| Critical (die) illness | 1.067% | 0.351% | 0.807% | 0.659% | 0.033% | 0.011% | 0.025% | 0.020% |
| **Clinical outcomes** | **Group A_12-17yrs** | **Group B_12-17yrs** | **Group C_12-17yrs** | **Group D_12-17yrs** | **Group A_5-11yrs** | **Group B_5-11yrs** | **Group C_5-11yrs** | **Group D_5-11yrs** |
| Asymptomatic infection | 33.00% | 33.17% | 33.06% | 33.10% | 33.00% | 33.17% | 33.06% | 33.10% |
| Mild/Moderate illness | 66.23% | 66.58% | 66.36% | 66.43% | 66.23% | 66.58% | 66.36% | 66.43% |
| Severe illness | 0.623% | 0.205% | 0.471% | 0.385% | 0.684% | 0.225% | 0.517% | 0.423% |
| Critical (recover) illness | 0.138% | 0.045% | 0.104% | 0.085% | 0.081% | 0.027% | 0.061% | 0.050% |
| Critical (die) illness | 0.009% | 0.003% | 0.006% | 0.005% | 0.005% | 0.002% | 0.003% | 0.003% |

#### 1.5 Direct medical cost

We collected the direct medical costs of COVID-19 disease in the US from published literature and Medicare Administrative Contractor report[15-17]. The cost of PCR tests and rapid antigen self-test for COVID-19 infection was estimated to be $51 and $11 per person, respectively. In addition, we collected cost per outpatient visit, general hospitalisation and ICU admission and the duration of each disease stage[18]. The cost of medical services varied across clinical disease stages, and we calculated the total direct medical cost of COVID-19 cases with varied severity by multiplying the unit cost of the medical services by the duration of each disease stage.

| Clinical outcomes | Self-tests cost (per person, $) | PCR test cost (per person, $) | Outpatient cost (per visit, $) | Hospitalization cost (per person, $) | | | ICU cost (per person, $) | | | Total direct medical cost (per person, $) |
|---|---|---|---|---|---|---|---|---|---|---|
| | | | | length of stay | per day cost | total cost | length of stay | per day cost | total cost | |
| Asymptomatic infection | 11 | 0 | 0 | 0 | 2364 | 0 | 0 | 3111 | 0 | 11 |
| Mild/Moderate illness | 11 | 51 | 164 | 0 | 2364 | 0 | 0 | 3111 | 0 | 226 |
| Severe illness | 11 | 51 | 164 | 9.2 | 2364 | 21752 | 0 | 3111 | 0 | 21978 |
| Critical (die) illness | 11 | 51 | 164 | 4.2 | 2364 | 9930 | 7.1 | 3111 | 22085 | 32241 |
| Critical (recover) illness | 11 | 51 | 164 | 10 (4.2+5.8) | 2364 | 23643 | 7.1 | 3111 | 22085 | 45954 |

#### 1.6 Health state utilities

We collected utility scores for COVID-19 patients from the disutility weights of severe lower respiratory tract infection[19,20] and the estimates of pricing models for COVID-19 treatments published by the Institute for Clinical and Economic Review[21]. We calculated the average utility from two sources and adopted their lowest and highest bounds[22].

| Health states | Disability weight 1 | Disability weight 2 | **Average utility** |
|---|---|---|---|
| In asymptomatic state | — | — | **1** |
| In mild/moderate state | — | — | **1** |
| In severe state | 0.13 (0.09, 0.19) | 0.30 | **0.785 (0.700, 0.910)** |
| In critical state | 0.41 (0.27, 0.56) | 0.50-0.60 | **0.520 (0.400, 0.730)** |
| In recuperable state | — | 0.19 | **0.905 (0.810, 1.000)** |
| In dead state | 0 | 0 | **0** |

#### 1.7 Sensitivity analyses

We extensively explored the impact of model parameters on the baseline results with univariate and probabilistic sensitivity analyses (PSA). Univariate sensitivity analysis is for each of 17 parameters and varying one parameter at one time in their range. PSA allows variation of all 17 parameters together at one time in their distribution with random sampling.

| No | Name | Distribution | Reference |
|---|---|---|---|
| 1 | Short-term booster VE for preventing non-BA.1 infection | Triangular (0.2611, 0.3545, 0.4362) | Appendix 1.2 |
| 2 | Short-term booster VE for preventing severe disease | Triangular (0.7276, 0.7876, 0.8343) | Appendix 1.2 |
| 3 | Long-term booster VE for preventing non-BA.1 infection | Triangular (0.0000, 0.0000, 0.0765) | Appendix 1.2 |
| 4 | Loge-term booster VE for preventing severe disease | Triangular (0.0960, 0.2441, 0.3679) | Appendix 1.2 |
| 5 | 2nd booster VE for preventing non-BA.1 infection, compared with no vaccination | Triangular (0.2411, 0.4006, 0.5266) | Appendix 1.2 |
| 6 | 2nd booster VE for preventing severe disease, compared with no vaccination | Triangular (0.4549, 0.6296, 0.7484) | Appendix 1.2 |
| 7 | Decrease in direct medical cost (%) | Uniform (-50, 50) | Assumed |
| 8 | Increase in vaccination cost (%) | Uniform (-50, 50) | Assumed |
| 9 | Utility of recuperable disease stage | Triangular (0.8100, 0.9050, 1.0000) | Appendix 1.6 |
| 10 | Utility of severe disease stage | Triangular (0.7000, 0.7850, 0.9100) | Appendix 1.6 |
| 11 | Utility of critical disease stage | Triangular (0.4000, 0.5200, 0.7300) | Appendix 1.6 |
| 12 | The discount rate | Uniform (0, 0.06) | [23] |
| 13 | Transmission coefficient (beta_0-4yrs) | Triangular (0.0043, 0.0054, 0.0065) | Appendix 1.3 and assumed |
| 14 | Transmission coefficient (beta_5-11) | Triangular (0.0031, 0.0039, 0.0047) | Appendix 1.3 and assumed |
| 15 | Transmission coefficient (beta_12-17) | Triangular (0.0020, 0.0024, 0.0029) | Appendix 1.3 and assumed |
| 16 | Transmission coefficient (beta_18-49) | Triangular (0.0027, 0.0034, 0.0041) | Appendix 1.3 and assumed |
| 17 | Transmission coefficient (beta_50+) | Triangular (0.0026, 0.0033, 0.0039) | Appendix 1.3 and assumed |

### 1.8 The impact of contact matrix on the results

To examine the impact of contact matrix on the findings, we assumed all age groups interact only within their peers. We re-calibrate the model to the number of death cases and obtain the following contact matrix.

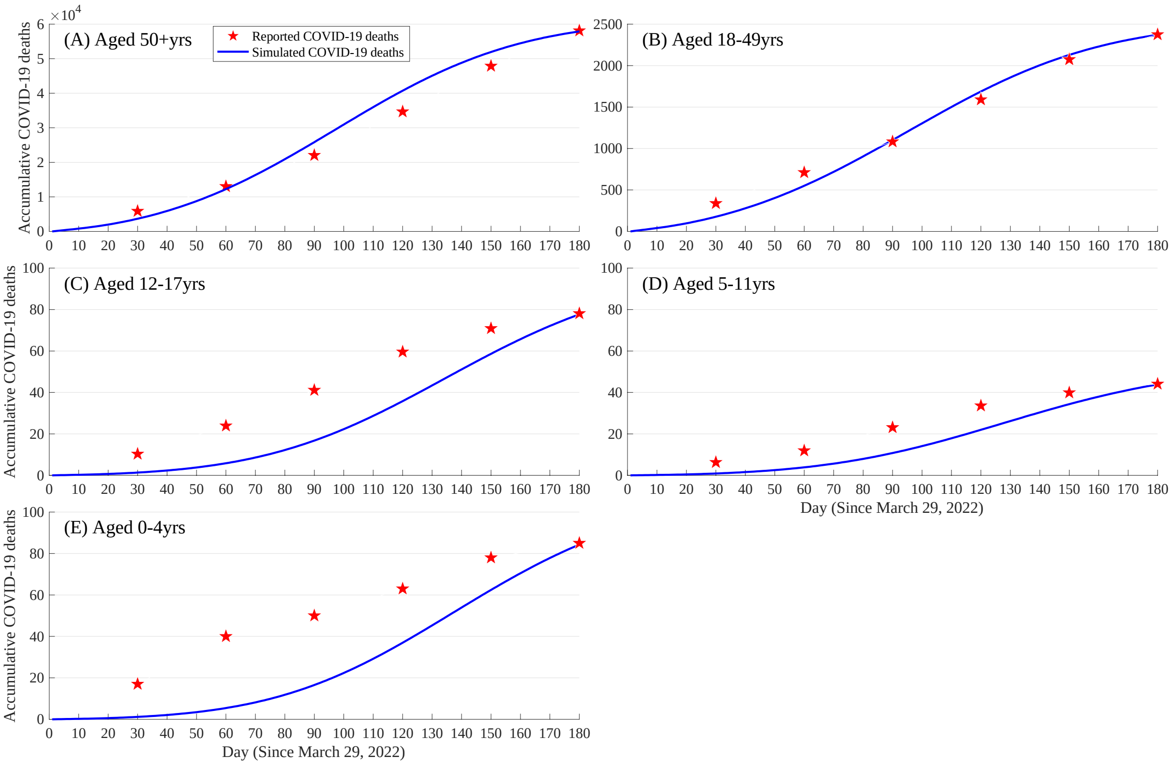


**Figure S4.** Model re-calibration for COVID-19 related deaths by age groups in the US.

The new contact metric in the US is obtained from the polished literature as follow.

| Susceptible \ Infections | 0-4 | 5-11 | 12-17 | 18-49 | ≥50 |
|---|---|---|---|---|---|
| 0-4 | 18.48 | 0 | 0 | 0 | 0 |
| 5-11 | 0 | 24.69 | 0 | 0 | 0 |
| 12-17 | 0 | 0 | 29.46 | 0 | 0 |
| 18-49 | 0 | 0 | 0 | 12.73 | 0 |
| ≥50 | 0 | 0 | 0 | 0 | 10.61 |

**Table S1 Model input parameters, values, and sources.**

| Model parameters | Values | Reference |
|---|---|---|
| **Epidemiological parameters** | | |
| Duration of clinical disease stages of COVID-19 (days) | | |
| Incubation | 5.2 (4.1, 7.0) | |
| Asymptomatic infection | 6.0 (2.0, 12.0) | |
| Mild/Moderate illness | 10.0 | |
| Severe illness | | |
| In mild/moderate state | 6.5 | |
| In severe state | 9.2 | |
| Critical (recover) illness | | |
| In mild/moderate state | 3.0 | |
| In severe state | 4.2 | |
| In critical state | 7.1 | |
| In recuperable state | 5.8 | |
| Critical (die) illness | | |
| In mild/moderate state | 3.0 | |
| In severe state | 4.2 | |
| In critical state | 7.1 | |
| **Costing parameters** | | Appendix 1.5 |
| Per PCR test for COVID-19 infection, $ | 51 | |
| Per rapid antigen self-test for COVID-19 infection, $ | 11 | |
| Per general practitioner (GP) consultation, $ | 164 | |
| Per day of general hospitalization, $ | 2364 | |
| Per day of ICU hospitalization, $ | 3111 | |
| Per dose of COVID-19 vaccine booster, $ | 17.25 | |
| Per dose of vaccine administration, $ | 17.10 | |
| Direct medical cost of varied COVID-19 severity, $ | | |
| Asymptomatic infection | 11 | |
| Mild/Moderate illness | 226 | |
| Severe illness | 21978 | |
| Critical (recover) illness | 32241 | |
| Critical (die) illness | 45854 | |
| **Life quality parameters** | | |
| Utility weight of clinical disease progression severity | | Appendix 1.6 |
| In asymptomatic state | 1 | |
| In mild/moderate state | 1 | |
| In severe state | 0.785 (0.700, 0.910) | |
| In critical state | 0.520 (0.400, 0.730) | |
| In recuperable state | 0.905 (0.810, 1.000) | |
| In dead state | 0 | |
| Discount rate, per year | 3% (0%, 6%) | [23] |
